# Day case versus inpatient total shoulder arthroplasty: A retrospective cohort study and cost-effectiveness analysis

**DOI:** 10.1101/2020.03.24.20036558

**Authors:** Aditya Borakati, Asad Ali, Chetana Nagaraj, Srinivas Gadikoppula, Michael Kurer

## Abstract

**Background:** Day case total shoulder arthroplasty (TSA) is a novel approach, not widely practiced in Europe. We conducted a retrospective cohort study of patients comparing elective day case and inpatient TSAs in our UK centre.

**Aim:** To evaluate the efficacy and cost-effectiveness of day case total shoulder arthroplasty (TSA) compared to standard inpatient total shoulder arthroplasty.

**Methods:** All patients undergoing TSA between January 2017 and July 2018 were included. Outcome measures were: change in abduction and extension 3 months postoperatively; 30 day postoperative adverse events and re-admissions in day case and inpatient groups. We also conducted an economic evaluation of outpatient arthroplasty. Multivariate linear and logistic regression were used to adjust for demographic and operative covariates.

**Results:** 59 patients were included, 18 day cases and 41 inpatients. There were no adverse events or re-admissions at 30 days postoperatively in either group. There were no significant differences in adjusted flexion (mean difference 16.4°; 95% CI -17.6° to 50.5°, p=0.337) or abduction (mean difference 13.2° 95% CI; -18.4° to 44.9°, p=0.405) postoperatively between groups. Median savings with outpatient arthroplasty were GBP 529 (IQR 247.33 to 789, p<0.0001).

**Conclusion:** Day case TSA is a safe, effective procedure, with significant cost benefit. Wider use may be warranted in the UK and beyond, with potential for significant cost savings and improved efficiency.

**Core tip:** In this article we show that day case total shoulder arthroplasty is a feasible, safe and effective alternative to inpatient admission for the same procedure, with an associated average cost saving of GBP 529.

## Introduction

Total shoulder arthroplasty is well established as a safe, effective treatment for multiple pathologies, including osteoarthritis, rotator cuff arthropathy and complex fractures of the humerus[1,2].

Typically, it is performed under general anaesthesia with an overnight stay for administration of analgesia intravenously. Increasingly, however, the procedure is performed on an outpatient (or day case) basis. This has been facilitated through the use of continuous nerve blockade with portable infusion devices for ambulatory analgesia outside of the hospital setting[3].

The latter approach has been widely adopted across the United States[4,5], although uptake has been slow elsewhere, notably in the United Kingdom and Europe, with limited literature available. There is a significant potential for improved cost effectiveness and throughput efficiency with an outpatient approach. We have therefore conducted this retrospective cohort study in our UK centre with the aim of comparing the clinical and cost efficacy of traditional inpatient total shoulder arthroplasty with outpatient regimens in this setting. We hypothesised that our outpatient protocol would have equivalent efficacy to inpatient protocols with a significantly lower cost.

## Materials and Methods

### Inclusion Criteria

All adult (18+) patients undergoing elective total shoulder arthroplasty at North Middlesex University Hospital, London, UK between January 2017 and July 2018 were included. Both anatomical and reverse arthroplasties were included.

### Exclusion Criteria

Patients undergoing arthroplasty for an acute traumatic indication, such as fracture, were excluded.

### Perioperative Procedure

Prior to selection for surgery all patients underwent clinical assessment of mobility with range of motion recorded. All patients selected for surgery underwent anaesthetic pre-assessment and were identified based on pre-morbid status (no severe cardiorespiratory co-morbidities) for suitability for an outpatient procedure. Patients must have had a friend or family member staying with them for 24 hours postoperatively, speak English, be contactable by telephone and have the family member willing to be trained to remove the analgesic catheter. Patients eligible for outpatient arthroplasty only underwent day case analgesic procedure and same day discharge dependent on staffing availability.

All patients were admitted on the day of surgery and operated on under general anaesthesia. Patients selected for a day case procedure were given a continuous intrascalene analgesic infusion following the surgery and discharged the same day, with the infusion pump in situ. This was then removed at day 3 postoperatively in the community by family members or friends of the patient who were given written instructions on removal. Patients were followed up daily over telephone by specialist pain nurses until removal of the infusion catheter. Those with planned inpatient stays were given strong opiate analgesia and were admitted until this could be weaned off with appropriate mobilisation as assessed by physiotherapy. Patients were followed up at 3 months post-operatively to assess mobility and to assess imaging to ensure appropriate prosthesis placement.

### Data Collection

Data was collected retrospectively, with patients identified from prospectively recorded theatre logs and case note retrieval. Data collected included demographic information such as age and gender, co-morbidities and indication for procedure, operative information and post-operative complications and range of motion at 3 months as assessed in clinic. Patients were stratified into day case and inpatient groups depending on the pre-operative plan for admission or not.

### Outcomes

Primary outcome measures were mean increase in active flexion and abduction at 3 months post-operatively. Secondary outcomes were postoperative complication and re-admission rates at 30 days.

### Economic analysis

The difference in costs between day case and inpatient procedures was calculated using the cost of catheter insertion, analgesic infusion costs and removal of the catheter for the outpatient group. For the inpatient group the median length of stay was used to calculate cost of inpatient nursing care. All other costs of care were assumed equal in both groups.

The median cost difference between inpatient and outpatient groups was calculated and statistical significance assessed using the Mann Whitney U test.

### Statistical Analysis

Descriptive data are presented as means with standard deviation or medians with interquartile ranges dependent on the normality of the data as appropriate. Normality was assessed by visual inspection of histograms and QQ plots and subsequent statistical testing was directed by this assessment. Statistical significance in terms of demographic and operative differences between day case and inpatient groups was calculated using Fisher’s exact or Student’s two-tailed t-tests as appropriate.

Difference in mean abduction and flexion between inpatient and outpatient groups was assessed using Student’s two-tailed t-test. The threshold for statistical significance was set with α < 0.05.

Multivariate linear and logistic regression analysis was used to adjust for demographic and operative covariates (including age, gender, side, anatomic versus reverse and indication of procedure) in the primary outcomes, following univariate analysis.

Statistical analysis was conducted using Microsoft R Open 3.5.1[6] (Microsoft Corp., Redmond, WA, USA) with tidyverse[7], desctools[8], finalfit[9], tableone[10] and lubridate[11] packages.

Following the main analysis we later decided (post-hoc) to conduct an analysis of the power of our sample to detect a clinically significant difference (agreed by consensus of the study team) of 30 degrees of abduction between outpatient and inpatient groups. This analysis used the variance in abduction following total shoulder arthroplasty at 6 months as reported by Ramzjou et al[12].

### Ethical Approval

This study was registered with the local clinical governance department at North Middlesex University Hospital as a service evaluation. No formal research ethics committee opinion was sought for this retrospective study with no alteration to patient care.

### Reporting Guidelines

This study is reported according to the STROBE guidelines for observational studies[13].

## Results

59 eligible patients were identified in the study period, of these, 18 were planned day cases and 41 planned inpatient stays. The characteristics of the study population are summarised in table 1, note-all the planned day case patients were discharged on the day of surgery as expected and all inpatients were admitted overnight as planned. Inpatients had a median stay of 2 days and 6 hours (IQR 28 to 78 hours). There were no significant differences in day case and inpatient groups at baseline.

**Table 1.**
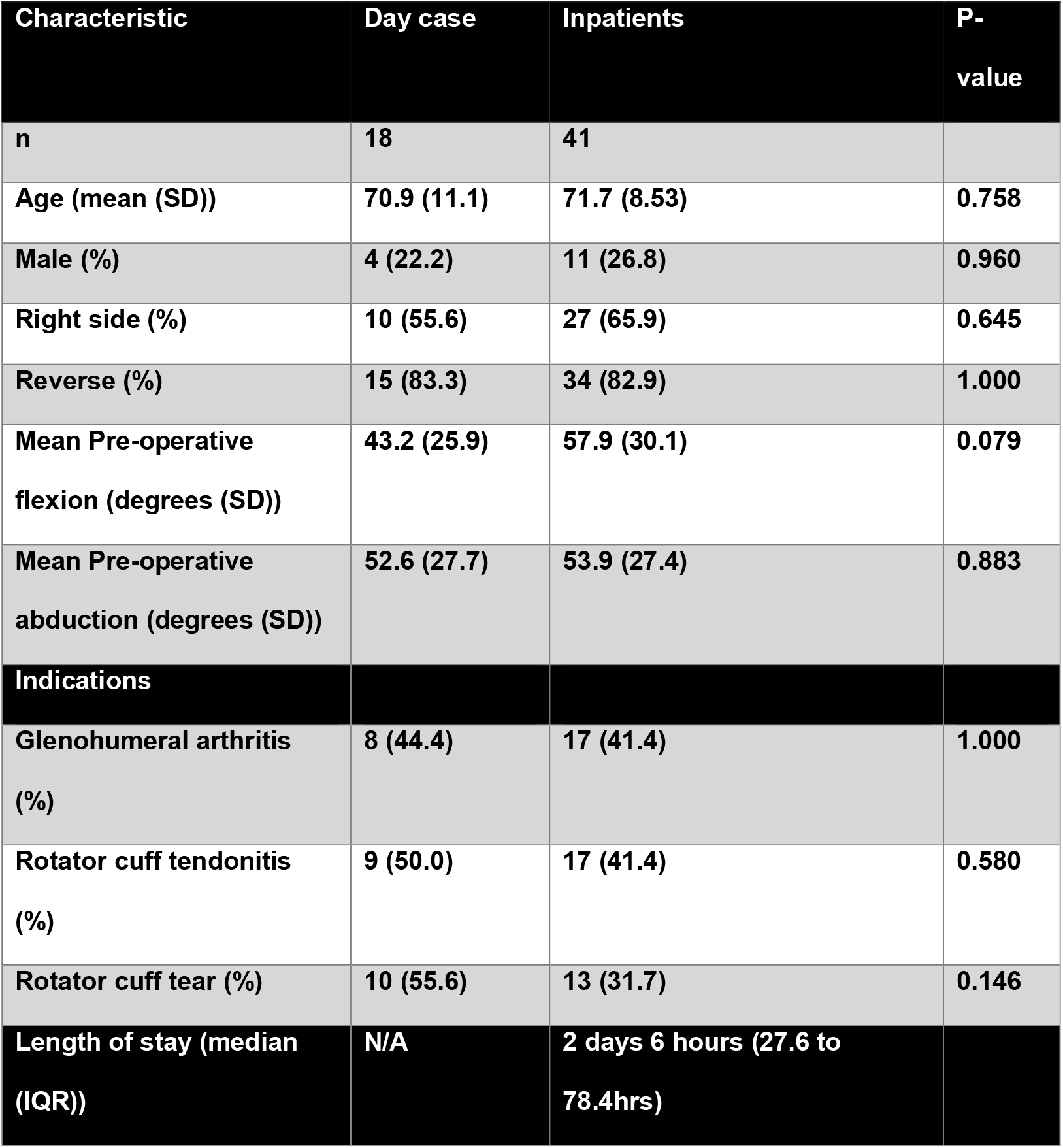
Summary characteristics of study population (note for indications, patients may have one or more pathologies)

| Characteristic | Day case | Inpatients | P-value |
| --- | --- | --- | --- |
| n | 18 | 41 |  |
| Age (mean (SD)) | 70.9 (11.1) | 71.7 (8.53) | 0.758 |
| Male (%) | 4 (22.2) | 11 (26.8) | 0.960 |
| Right side (%) | 10 (55.6) | 27 (65.9) | 0.645 |
| Reverse (%) | 15 (83.3) | 34 (82.9) | 1.000 |
| Mean Pre-operative flexion (degrees (SD)) | 43.2 (25.9) | 57.9 (30.1) | 0.079 |
| Mean Pre-operative abduction (degrees (SD)) | 52.6 (27.7) | 53.9 (27.4) | 0.883 |
| **Indications** |  |  |  |
| Glenohumeral arthritis (%) | 8 (44.4) | 17 (41.4) | 1.000 |
| Rotator cuff tendonitis (%) | 9 (50.0) | 17 (41.4) | 0.580 |
| Rotator cuff tear (%) | 10 (55.6) | 13 (31.7) | 0.146 |
| Length of stay (median (IQR)) | N/A | 2 days 6 hours (27.6 to 78.4hrs) |  |

Unadjusted analysis (table 2) showed no significant difference between groups for the increase in range of motion post-operatively. Univariate analysis of the demographic and operative covariates, alongside admission status is displayed in table 3, there were no significant associations between the primary outcomes and the explanatory variables.

**Table 2.** Unadjusted changes in flexion, abduction and discharge rates at 3 month post-operatively in day case and inpatient groups

| Characteristic | Day case | Inpatients | P-value |
| --- | --- | --- | --- |
| n | 18 | 41 | - |
| Post-op change in Flexion (mean degrees (SD)) | 32.7 (52.1) | 48.6 (53.7) | 0.325 |
| Post-op change in Abduction (mean degrees (SD)) | 42.5 (47.8) | 33.2 (58.7) | 0.528 |

**Table 3.** Uni- and multi-variate analyses of postoperative range of motion difference

| Explanatory variable | Univariate Flexion (degrees) |  | Multivariate Flexion (degrees) |  |
| --- | --- | --- | --- | --- |
|  | Mean Increase (95% CI) | p-value | Mean Increase (95% CI) | p-value |
| Day case (versus inpatient) | 15.9 (-16.2 to 48.0) | 0.325 | 16.4 (-17.6 to 50.5) | 0.337 |
| Age (increasing) | 0.50 (-1.11 to 2.12) | 0.534 | 1.71 (-0.45 to 3.88) | 0.118 |
| Gender (male) | -3.30 (-30.9 to 37.5) | 0.848 | -11.7 (-25.4 to 48.9) | 0.530 |
| Laterality (right) | -29.3 (-59.1 to 0.56) | 0.054 | -36.7 (-70.4 to -3.08) | 0.033 |
| Reverse/ Anatomical | -1.76 (-41.5 to 38.0) | 0.930 | -16.4 (-50.4 to 17.5) | 0.337 |
| Glenohumeral arthritis | -13.8 (-43.6 to 15.9) | 0.357 | -18.8 (-52.4 to 14.8) | 0.266 |
| Cuff tendonitis | -0.08 (-30.11 to 29.96) | 0.996 | -9.93 (-48.1 to 28.2) | 0.603 |
| Cuff tear | 8.66 (-21.83 to 39.15) | 0.572 | -5.67 (-43.1 to 31.7) | 0.762 |
| Explanatory variable | Univariate Abduction (degrees) |  | Multivariate Abduction (degrees) |  |
|  | Mean Increase (95% CI) | p-value | Mean Increase (95% CI) | p-value |
| Day case (versus inpatient) | 9.33 (-20.1 to 38.8) | 0.528 | 13.2 (-18.4 to 44.9) | 0.405 |
| Age (increasing) | 1.05 (-0.40 to 2.50) | 0.152 | 1.97 (-0.03 to 3.98) | 0.054 |
| Gender (male) | 5.34 (-25.9 to 36.6) | 0.733 | 7.68 (-26.8 to 42.2) | 0.657 |
| Laterality (right) | -7.44 (-35.5 to 20.6) | 0.598 | -17.3 (-48.6 to 13.9) | 0.271 |
| Reverse/ Anatomical | 9.05 (-27.1 to 45.2) | 0.618 | -9.10 (-56.9 to 38.7) | 0.704 |
| Glenohumeral arthritis | -9.04 (-36.3 to 18.3) | 0.510 | -17.5 (-48.6 to 13.7) | 0.266 |
| Cuff tendonitis | -7.66 (-35.0 to 19.7) | 0.577 | -20.2 (-55.6 to 15.2) | 0.256 |
| Cuff tear | 6.53 (-21.3 to 34.4) | 0.641 | -7.69 (-42.4 to 27.0) | 0.658 |

Following adjustment for all covariates (table 3 and figure 1)-there was no significant difference between day case and inpatient groups for change in flexion (mean difference 16.4°; 95% CI -17.6° to 50.5°, p=0.337) and abduction postoperatively (mean difference 13.2°; 95% CI; -18.4° to 44.9°, p=0.405).

**Figure 1.**
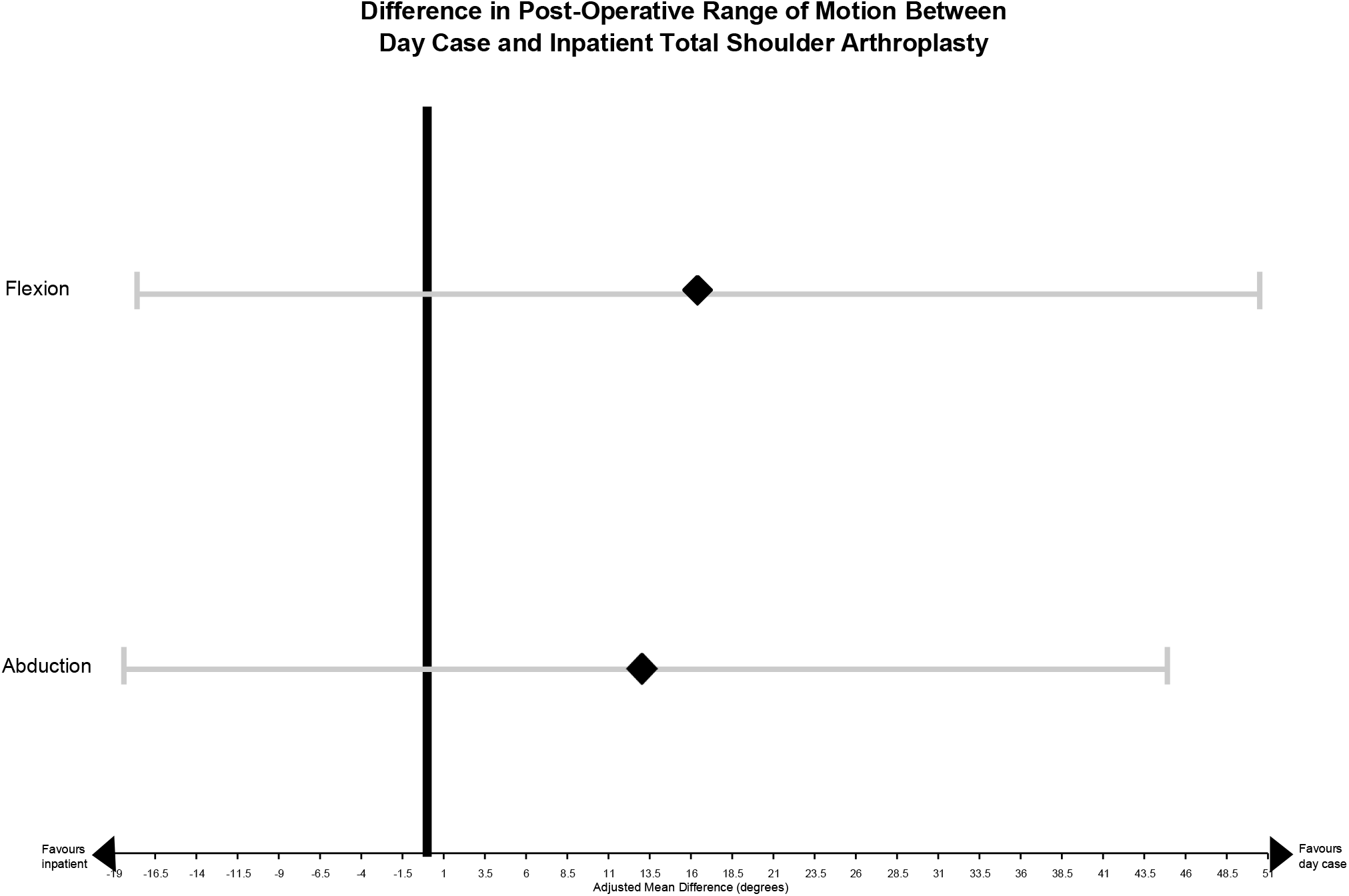
Adjusted Mean Difference in postoperative flexion and extension between day case and inpatient groups at 3 months. Bars represent 95% confidence intervals.

There were no adverse events related to surgery in both groups and no re-admissions following discharge in either group.

### Economic analysis

Mean cost of admission was £260 per day. This gave a median cost of admission for the inpatient group of £585 (IQR 303.33 to 845). Cost of catheter insertion, infusion and removal was £56. Cost of the analgesia catheter was the same for all outpatients. It is assumed all other variables are the same between inpatient and outpatient groups.

The median savings of outpatient arthroplasty were therefore £529 (IQR 247.33 to 789.00, p<0.0001).

### Power calculation

Post-hoc power calculations showed with this sample size, there was a 0.861 power to detect a 30 degree difference in abduction using the variance reported by Razmjou et al. at 6 months[12].

## Discussion

This study is the only published experience of outpatient total shoulder arthroplasty in the UK. We show the non-inferiority of day case total shoulder replacement with inpatient total shoulder arthroplasty in terms of range of motion and adverse events post-operatively, at lower cost.

Our results mirror those reported elsewhere. Ilfield et al showed similar range of motion outcomes in inpatient and outpatient groups in their initial pilot study with a single intrascalene block post-operatively in 2005[3]. In their follow up randomised trial they further showed patients who did not have the block, had a lower range of motion initially after the operation as would be expected, despite high doses of intravenous opioids[14].

Recently, Bean and colleagues in their similarly sized retrospective study in the United States, showed reduced 90-day complication rates with outpatient arthroplasty and fewer visits to emergency departments following discharge compared to their inpatient comparator[15]. In this study, patients were excluded from receiving outpatient surgery if they had a number of comorbidities, such as cardiopulmonary disease. Leroux and Basques et al. in their population level studies in the US also confirmed no increased adverse events or re-admissions in those undergoing outpatient surgery, even after adjusting for pre-existing co-morbidities and demographic factors such as age[4,5].

The cost benefit of outpatient shoulder arthroplasty has already been modelled in the US, where savings were estimated between 747 and 15507 USD per patient, with a base case of 5594 USD[16]. Our findings match the lower end of these estimates, however, we have only evaluated two key variables for cost differential in our analysis. Additional analgesics, blood tests and physiotherapy costs are likely to increase the relative cost of standard inpatient arthroplasty and we have not accounted for these. Further, admission costs in the US are known to be markedly higher than the UK[17].

In addition to direct costs, the potential for increased throughput and reduction of bed use is significant, particularly in the context of the NHS where there are significant waiting times and targets to be met.

Our study is also relatively unique in having solely remote follow up of patients with the infusion catheter. Many centres have specialist community nursing teams to facilitate the care and monitoring of the analgesic infusion whilst in the community. Our study shows that it is safe to monitor these patients remotely, while alleviating the need for specialist community nursing resources and training.

In the UK the only published experience of nerve block infusions for ambulatory shoulder surgery was a successful pilot of 10 patients, which showed good analgesia[18].

### Strengths

Our study is relatively unique outside of the US and has a comparator group of inpatients unlike many similar studies. We have further looked at functional outcomes in terms of range of motion in comparison to inpatient surgery unlike much of the previous literature. This study also contains the only cost analysis of outpatient shoulder arthroplasty outside the US. We believe our study has greater applicability to the publicly provided health systems found in the UK and Europe, both in terms of demographic similarity and patient pathways.

Further, the statistical analysis we have conducted is robust and the multivariate analysis we have conducted includes several covariates which have been adjusted for. Finally, we have conducted a power calculation, which whilst post-hoc, shows that this study may be appropriately powered and can be used to inform future studies.

## Limitations

We have not been able to collect data on co-morbidities to adjust for this as a covariate for post-operative results. Further we have not collected data on pain scores or satisfaction in the long term, which are key factors in the success of the operation. Further, although we have collected data on the two key movements (extension and abduction) ideally a formalised functional assessment such as the Oxford Shoulder Score[19] should be used. However, in theory, as the operative procedure is identical in both inpatient and outpatient groups, there should be no long term differences in pain or function with the outpatient method, which only alters the post-operative analgesia modality.

For the cost analysis we did not look at all possible costs associated with each procedure and their respective pathways but only the key differentiators, the cost of the catheter and infusion for outpatients and the cost of admission of inpatients (the latter only includes nursing costs). Preferrably, alternative costs such as anaesthetic and recovery times, pain nurse telephone follow up, inpatient physiotherapy, pain nurse and medication costs should be included.

Ideally, an appropriately powered, randomized controlled trial with long term follow up comparing patients undergoing outpatient arthroplasty with a continuous nerve block versus the traditional inpatient group is needed, as yet, no such trial has been conducted. This trial should randomise patients regardless of pre-existing co-morbidities and assess postoperative complication rates, re-admission rates as well as postoperative function and overall service discharge rate. A comprehensive cost analysis using NHS tariff prices is essential following this to ensure translation in our setting.

The criterion regarding appropriate selection of patients is important, as there is much debate around this, in shoulder but also hip and knee arthroplasty, where outpatient surgery has been more extensively studied[20]. Meneghini et al. have recommended a scoring system to facilitate selection of suitable patients for day case arthroplasty, however, this only considered hip and knee arthroplasties[21]. Ideally, a similar scoring system or set of criteria to identify eligible patients for day case shoulder arthroplasties is needed.

Further, it may be advantageous to use regional anaesthesia only as opposed to a general anaesthetic to further reduce recovery times and potentially to extend surgery to those unfit for general anaesthesia. This has been employed in arthroscopic procedures and some more minor shoulder surgeries such as rotator cuff repair, but is rarely used for total shoulder arthroplasty due to inadequate analgesia. Development of newer techniques such as continuous intrascalene blocks and alternative blocks such as supraclavicular blocks may ameliorate this[22].

It must be noted that the peripheral nerve block techniques that facilitate day case shoulder arthroplasty are not without their disadvantages. The process of achieving regional nerve blockade takes significantly more time than induction of general anaesthesia[23]. With intrascalene blockades, complications include pneumothoraces, phrenic nerve palsies, transient or permanent neurological deficits including hoarseness of voice and Horner’s syndrome due to incorrect nerve blockade and, rarely, systemic toxicity such as myocardial depression. Overall, the risks of regional anaesthesia for surgery are still far lower and less severe than with general anaesthesia[24].In conclusion, elective total shoulder arthroplasty appears to be safe and effective when performed as a day case procedure in the UK, with lower costs, mirroring similar results reported in the USA. This suggests that these procedures should be performed more widely as a day case procedure in the UK and other countries with similar publicly funded health systems, in carefully selected patients, to reduce bed occupancy, improve efficiency and reduce costs. However, larger, more rigorous randomised controlled trials comparing the day case procedure with traditional inpatient regimes including robust cost-effectiveness analyses are needed.

## Article highlights

### Research background

Total shoulder arthroplasty is typically performed as an inpatient procedure with an overnight stay for adequate analgesia and observation. Advances in regional anaesthesia have enabled this major operation to be conducted as an outpatient procedure. The safety, efficacy and cost-effectiveness of the outpatient procedure are well established in the United States, but evidence and experience in the techniques are lacking elsewhere.

### Research motivation

Worldwide, there is significant scarcity in healthcare resources in terms of funding and bed capacity. These pressures are particularly serious in publicly funded health systems, such as that in the UK’s National Health Service, where we report our experience. Performing procedures such as total shoulder arthroplasty as outpatient procedures may reduce bed occupancy while obtaining significant cost benefits. The study was registered with our local clinical governance department as a service evaluation, no explicit patient consent was required for this study of anonymised retrospective data.

### Research objectives

We aimed to compare standard inpatient total shoulder arthroplasty with outpatient total shoulder arthroplasty. The primary outcomes were change in flexion and extension at 3 months postoperatively in each group. Adverse events, re-admission rates and cost analyses were also obtained.

### Research methods

We conducted a retrospective cohort study of all patients who underwent total shoulder arthroplasty at North Middlesex University Hospital, London, UK between January 2017 and July 2018. Both inpatient and outpatient surgical groups underwent general anaesthesia and the same operative procedures. The outpatient group had continuous intrascalene analgesic infusion catheters which were retained postoperatively and they were discharged on the day of surgery. These patients were followed up by telephone by specialist community pain nurses for 3 days postoperatively and the catheter removed by the patient in their home on day 3. Costs were calculated with median length of stay and admission costs in the inpatient group and catheter, infusion and community nursing costs in the outpatient group.

Between group differences were assessed using Student’s T-test or chi squared tests as appropriate. Multivariate linear and logistic regression was conducted to adjust for confounding variables.

### Research results

59 patients were included, 18 day cases and 41 inpatients. There were no adverse events or re-admissions at 30 days postoperatively in either group. There were no significant differences in adjusted flexion (mean difference 16.4°; 95% CI -17.6° to 50.5°, p=0.337) or abduction (mean difference 13.2° 95% CI; -18.4° to 44.9°, p=0.405) postoperatively between groups. Median savings with outpatient arthroplasty were GBP 529 (IQR 247.33 to 789, p<0.0001).

### Research conclusions

This study shows that outpatient total shoulder arthroplasty is a safe procedure with similar efficacy to traditional inpatient arthroplasty. We demonstrate significant cost savings with the outpatient procedure in our publicly funded, UK setting.

These findings suggest that outpatient total shoulder arthroplasty should replace traditional inpatient arthroplasty in suitable patients, in the UK and beyond, to save costs and relieve capacity.

### Research perspectives

Ideally an appropriately powered, randomised control trial comparing outpatient and inpatient procedures is required to evaluate the technique. Formal functional assessment with tools such as the Oxford Shoulder Score is also needed to accurately assess efficacy.

New methods of anaesthesia such as total regional anaesthesia with brachial plexus blockade need further study and may obviate the need for general anaesthesia and extend availability of surgery to those unfit for general anaesthesia.

Novel minimally invasive surgical techniques such as arthroscopic and robotic shoulder arthroplasty may also reduce pain and the need for inpatient admission.

### Biostatistics

The statistical methods of this manuscript were reviewed by Mr P Bassett of Statsconsultancy Ltd., Amersham, Bucks, UK. A letter approving the methods is enclosed as a supplementary file.

## Data Availability

Data is available on reasonable request to the corresponding author.

## Informed Consent

No informed consent was necessary for publication of anonymised results of this local service evaluation in line with local policies.

## Ethical Approval

This study was registered with the local clinical governance department at North Middlesex University Hospital as a service evaluation; no formal ethical approval was required.

## Guarantorship

**AB**

## Contributorship

AB contributed to the design, data collection, analysis and write up of the manuscript. CN and SG assisted with design of the study and data collection. AA and MK reviewed the manuscript.

This work has been presented (with more limited data including exclusion of cost effectiveness analysis) as a poster presentation at the Royal College Surgeons Orthopaedic Conference in Glasgow, UK on 17^th^ May 2019.

This work has been presented orally at the European Orthopaedic Research Society Conference in Maastricht, Netherlands on 4^th^ October 2019.

This work has not been published elsewhere, including in abstract form.

All authors have reviewed and approved the manuscript for publications. All authors believe that this manuscript represents honest work.

